# Leveraging patients’ longitudinal data to improve the Hospital One-year Mortality Risk

**DOI:** 10.1101/2024.06.21.24309191

**Authors:** Hakima Laribi, Nicolas Raymond, Ryeyan Taseen, Dan Poenaru, Martin Vallières

## Abstract

**Purpose:** Predicting medium-term survival after admission is necessary for identifying end-of-life patients who may benefit from goals of care (GOC) discussions. Considering that several patients have multiple hospital admissions, this study leverages patients’ longitudinal data and information collected routinely at admission to predict the Hospital One-year Mortality Risk.

**Methods:** We propose an Ensemble Long Short-term Memory neural network (ELSTM) to predict one-year mortality using patients’ longitudinal records. The model was evaluated: (i) with only predictors reported upon admission (AdmDemo); and (ii) also with diagnoses available later during patients’ stay (AdmDemoDx). Using records of 123,646 patients with 250,812 hospitalizations from 2011-2021, our dataset was split into a learning set (2011-2017) to compare models with and without longitudinal information using nested cross-validation, and a holdout set (2017-2021) to assess clinical utility towards GOC discussions.

**Results:** The ELSTM achieved a significant increase in predictive performance using longitudinal information (*p*-value < 0.05) for both the AdmDemo and AdmDemoDx predictors. For randomly selected hospitalizations in the holdout set, the ELSTM showed: (i) AUROCs of 0.83 (AdmDemo) and 0.87 (AdmDemoDx); and (ii) superior decision-making properties, notably with an increase in precision from 0.25 for the standard process to 0.28 (AdmDemo) and 0.36 (AdmDemoDx). Feature importance analysis confirmed that the utility of the longitudinal information increases with the number of patient hospitalizations.

**Conclusion:** Integrating patients’ longitudinal data provides better insights into the severity of illness and the overall patient condition, in particular when limited information is available during their stay.

## 1 Introduction

Estimating the life expectancy of patients helps identifying high-risk individuals and improve the quality of care they receive in hospital settings [1–3]. Unlike patients with cancer who receive palliative care in their final months of life, patients with other less predictable conditions are only referred for these services in their final weeks or days, if at all [4]. In Canada, despite common individual preference for most individuals to die in community and other home-like settings [5], 58% of those who died in 2015 were hospitalized more than once in their last year of life, and 61% died in hospital [6]. An early identification of these high-risk patients would allow important discussions with healthcare providers regarding end-of-life choices, to align their preferences with the care they receive [7]. Such discussions would enable goals-of-care (GOC) documentation, including code status orders (CSOs) clarifying essential preferences for life-supporting therapy [8, 9]. Early identification would also facilitate communication between clinicians and families regarding patients’ life trajectories, ensuring informed shared decision-making [10] and potentially reduce depression and grief [11]. However, a clear and timely prognostication of high-risk patients in hospital settings is time-consuming and therefore challenging for workload-burdened clinicians [12]. An accurate automated tool not requiring human involvement could initially flag these patients, lightening the work burden of the clinical team.

Several studies have investigated the ability of data available in Electronic Health Records (EHRs) to predict the mortality risk of patients, potentially driving an automated clinical decision support system. *van Walraven et al* [13, 14] introduced the Hospital One-year Mortality Risk (HOMR) score, representing the probability of death within one year of patient’s admission. The original model consisted of a logistic regression using post-discharge administrative data routinely collected upon admission, evaluated using Area Under the Receiver Operating Characteristic curve (AUROC). Their goal was to flag high-risk individuals and initiate end-of-life discussions with them to decide in favor or against potentially aggressive and invasive interventions. To operate in real-time settings, subsequent studies modified the HOMR score according to the availability of data in each hospital, and included only variables available immediately when patients were admitted [15, 16]. As a result, due to specific EHRs constraints, diagnostic codes were omitted from the predictors. More recently, *Taseen and Ethier* [9] explored the clinical utility of models predicting the HOMR score, in which they developed three random forest models based on variable sets available at different times during a patient’s admission. The authors compared the discriminative power of such models with previously established linear regression models and evaluated their clinical utility within their hospital setting.

Nevertheless, these studies did not include valuable longitudinal information present in patients’ records, as they focused on single visits and did not take into account the patient’s history from previous hospital admissions. This approach diverges from the clinical reality, where clinicians consistently consider the patient history before making a prognostic prediction for a given condition. Another approach proposed in previous work has been to incorporate broader covariates (e.g., medical disease codes, clinicians’ notes, social history) and aggregate patient information within and across admissions to predict their mortality risk in order to refer them for end-of-life care [17, 18]. However, these studies did not explicitly quantify the impact of integrating patient history in developing more accurate solutions. Moreover, these proposed models are more challenging in terms of data acquisition and are therefore less likely to be deployed in a clinical decision support system — unlike HOMR-based models that have already been clinically deployed [16] or are in the process of deployment [9].

In this work, we evaluated the benefits of integrating patients’ longitudinal data to improve the accuracy of the HOMR score. We built on the work of *Taseen and Ethier* [9] by reanalyzing the same data routinely collected during patients’ admissions while also integrating additional recent visits. To assess the benefits of a temporal EHR analysis, we developed and compared a Long Short-Term Memory-based ensemble model (ELSTM) that leverages patients’ longitudinal data, to baseline models that consider patients’ visits independently without including previous visits. Fig. 1 shows an overview of our study. We further analyzed the predictive power of our model in two different scenarios with different requirements of data access: (i) including only demographics and admission characteristics available on patient’s admission (“AdmDemo”), and (ii) adding also admission diagnoses and comoridity diagnoses available during patient’s hospitalization (“AdmDemoDx”). In an effort to better inform about the clinical utility of such models, we quantified the gains and losses of our ELSTM in terms of true and false positives as compared to standard human decision-making.

**Fig. 1.**
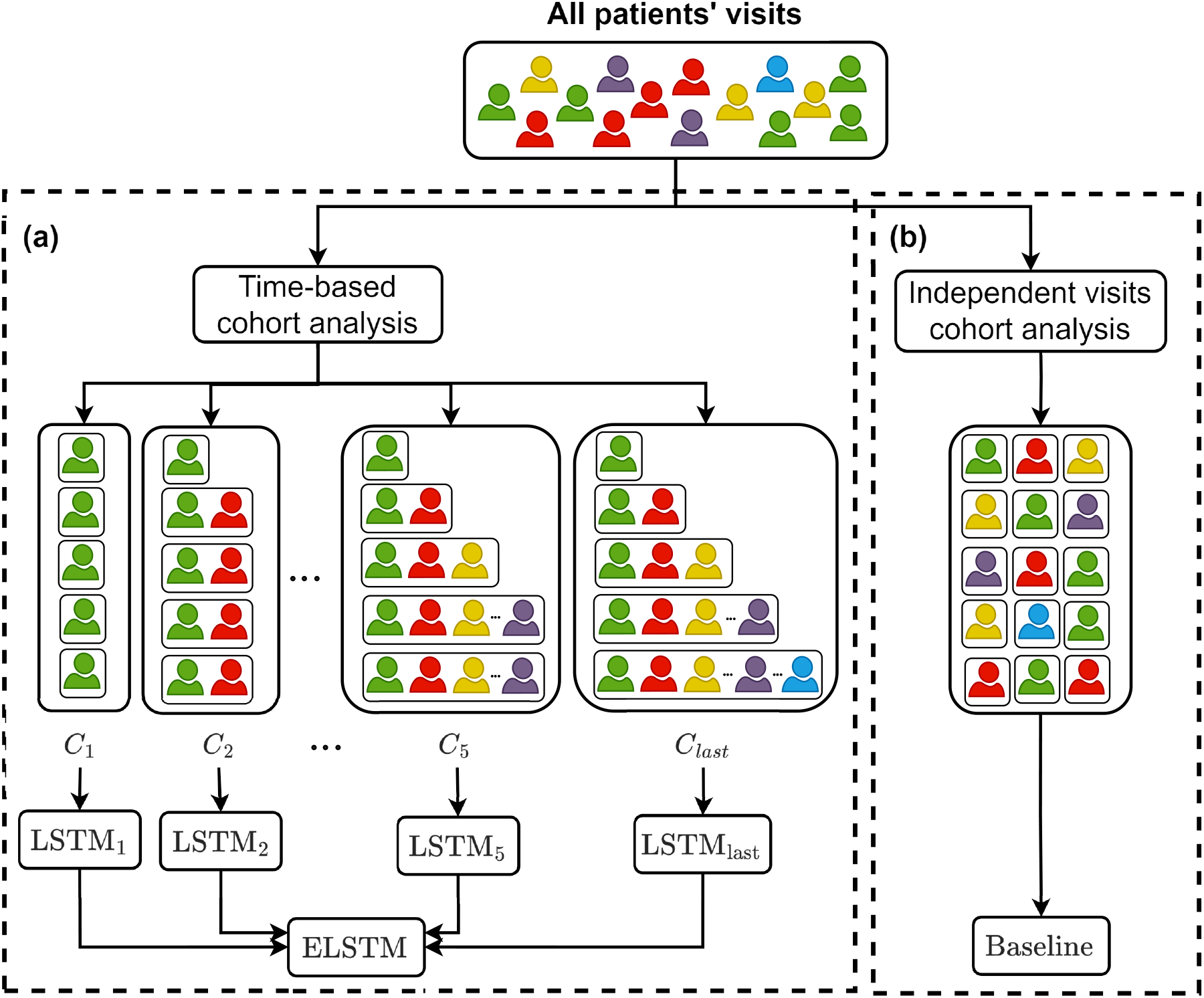
Study overview. (a) The ELSTM averages predictions of multiple LSTMs trained using different cohorts of the same patients. Each cohort includes the patient’s history up to a specific visit. (b) Baseline models consider patients visits independently

## 2 Materials and methods

### 2.1 Dataset

This retrospective study took place at an integrated university hospital network with 2 sites and 700 acute care beds in Sherbrooke, Quebec, Canada. Data was obtained from the institutional data warehouse, combining EHR and administrative information. The cohort included all adult patients admitted to a non-psychiatric service between July 1, 2011 and June 30, 2021, excluding admissions to infrequently admitting services (such as genetics) or admissions with a legal context (i.e. court-ordered). Mortality status was also extracted from the institutional data warehouse, which was sourced from the Quebec vital statistics registry. Institutional Review Board approval was obtained prior to data acquisition (Institutional Review Board of the CIUSSS de l’Estrie—CHUS Nagano #2022-4409). We followed the data extraction steps previously described by *Taseen and Ethier* [9] since we used the same source of data. Table 1 lists the predictors used for model comparisons. Comorbidity diagnoses from prior visits became accessible in the information system 6 months following a given visit, or only 2 weeks later for emergency department encounters.

**Table 1:**
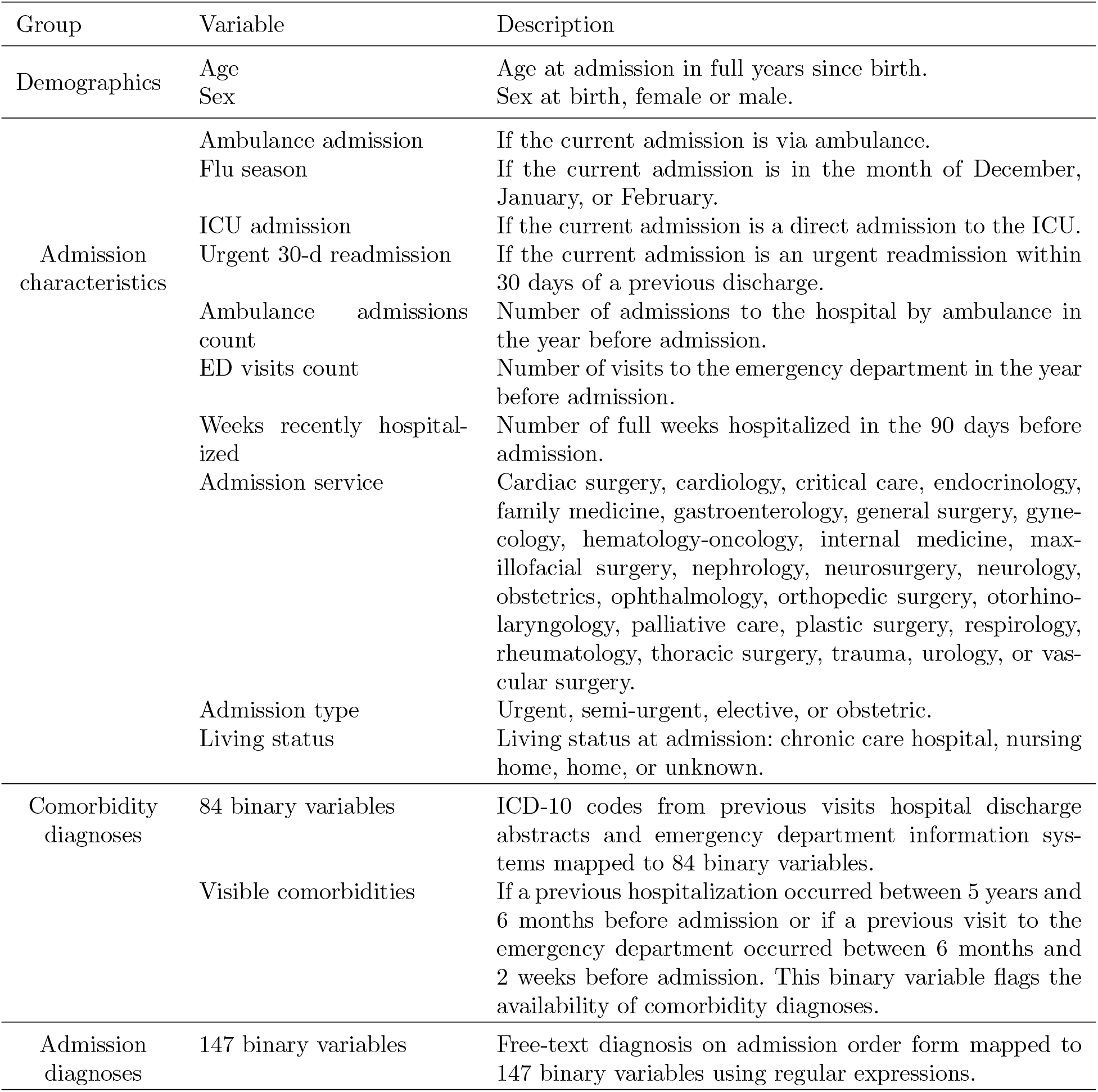
Covariates included in all predictive models as described in *Taseen and Ethier* [9]. AdmDemo predictors include only demographics and admission characteristics while AdmDemoDx predictors include demographics, admission characteristics and comorbidity and admission diagnoses.

Given the potential variations in data availability on admission across different hospital information systems, we explored the feasibility of early identification of high-risk patients in several scenarios. We evaluated two strategies with different data requirements: (i) “AdmDemo”, including only demographics and admission characteristics and, (ii) “AdmDemoDx” including demographics, admission characteristics, comorbidity diagnoses and admission diagnoses.

### 2.2 Ensemble Long Short-Term Memory neural network (ELSTM)

To evaluate the impact of incorporating a patient’s longitudinal health record for improving the HOMR score, we introduce an Ensemble Long Short-Term Memory neural network (ELSTM) that leverages information learned by multiple LSTMs trained at different stages of patients’ admissions to hospital (Fig. 1a). We base our ensemble model on an LSTM architecture [19] since recurrent neural networks can handle sequences of different lengths without extra padding. This is particularly relevant in our case where patients can have varying numbers of previous visits.

More formally, we define *C*_*k*_ as the temporal cohort including the visits sequence of each patient up to their *k*^*th*^ visit; if a patient has less than *k* visits, *C*_*k*_ includes all their visits. *C*_last_ denotes the cohort including the visits sequence of each patient up to their last visit available in our dataset. The formal definition of *C*_*k*_ is given by:

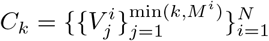

where *N* ∈ ℕ is the number of patients, *M*^*i*^ ∈ ℕ the number of visits for the *i*^*th*^ patient and 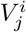 the *j*^*th*^ visit of the *i*^*th*^ patient.

During the training phase, we train multiple LSTMs on temporal cohorts including patients with varying numbers of visits. The goal is to capture diverse information at different stages of patients’ visit sequence. Each LSTM_k_ is trained using the temporal cohort *C*_*k*_ to aggregate a patient’s visit sequence and estimate their mortality risk at their last visit available in *C*_*k*_, with *k* ∈ {1, …, K} ∪ {last}. The ensemble model learns from multiple visits for each patient, while each LSTM_k_ is exclusively trained on a single visit sequence per patient. This setup guarantees that the training data for each LSTM_k_ are independent and identically distributed (iid). We set *K* = 5 given that only 5% of patients have more than 5 visits in our dataset. We chose not to restrict *C*_*k*_ to patients with only *k* visits in order to optimize each LSTM_k_ of the ensemble model on a larger set of data. Here, our assumption is that including patients with a full sequence of visits, even if the length was less than *k*, would make the distribution of training data more exhaustive and improve the model’s predictive performance.

In the testing phase, the ELSTM averages the predictions of all LSTMs trained with patients having at least *m* visits to make a prediction at the *m*^*th*^ visit of a patient, as follows:

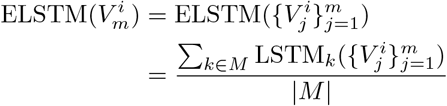

### 2.3 Experimental setup

#### 2.3.1 Baseline models

We conducted a comparative analysis of the ELSTM with two baseline models which do not use longitudinal data. The first model is the random forest (RF), as employed in prior work [9], using the scikit-learn wrapper [20] from skranger library^1^. The second model is a Basic LSTM (BLSTM) which does not consider previous information when making a prediction for a specific visit. Each LSTM-based model contains one single hidden layer followed by 2 fully connected layers and was implemented using the PyTorch library [21]. For a fair comparison, we added the previous visit count at each admission as a predictor to the baseline models.

#### 2.3.2 Experimental design

We used the experimental setup illustrated in Fig. 2 to evaluate the ELSTM and baseline models. The experiments are repeated for each group of predictors AdmDemo and AdmDemoDx. Following a similar approach to *Taseen and Ethier* [9], we temporally split the dataset into a *learning set*, including admissions from July 1, 2011, to June 30, 2017, and a *holdout set*, including admissions from July 1, 2017, to June 30, 2021. We excluded patients admitted before June 30, 2017 from the holdout set to prevent data leakage. This design aimed to simulate the evaluation of a model trained on all available patients data and tested on subsequently admitted patients. As patients are exclusively in one set at a time, temporal models have only aggregated previous visits occurring within the last six years prior to the current admission. To evaluate the final model’s clinical utility on the holdout set, we focused on the same population eligible for GOC discussions as in previous work [9]. Therefore, we excluded hospitalizations without an overnight stay from the holdout set, since there would not be enough time for a GOC discussion to occur. Additionally, we omitted admissions to the obstetrics service, where such discussions are considered inappropriate, and admissions to the palliative care service, where GOC discussions have already occurred and are therefore unnecessary at this stage.

**Fig. 2.**
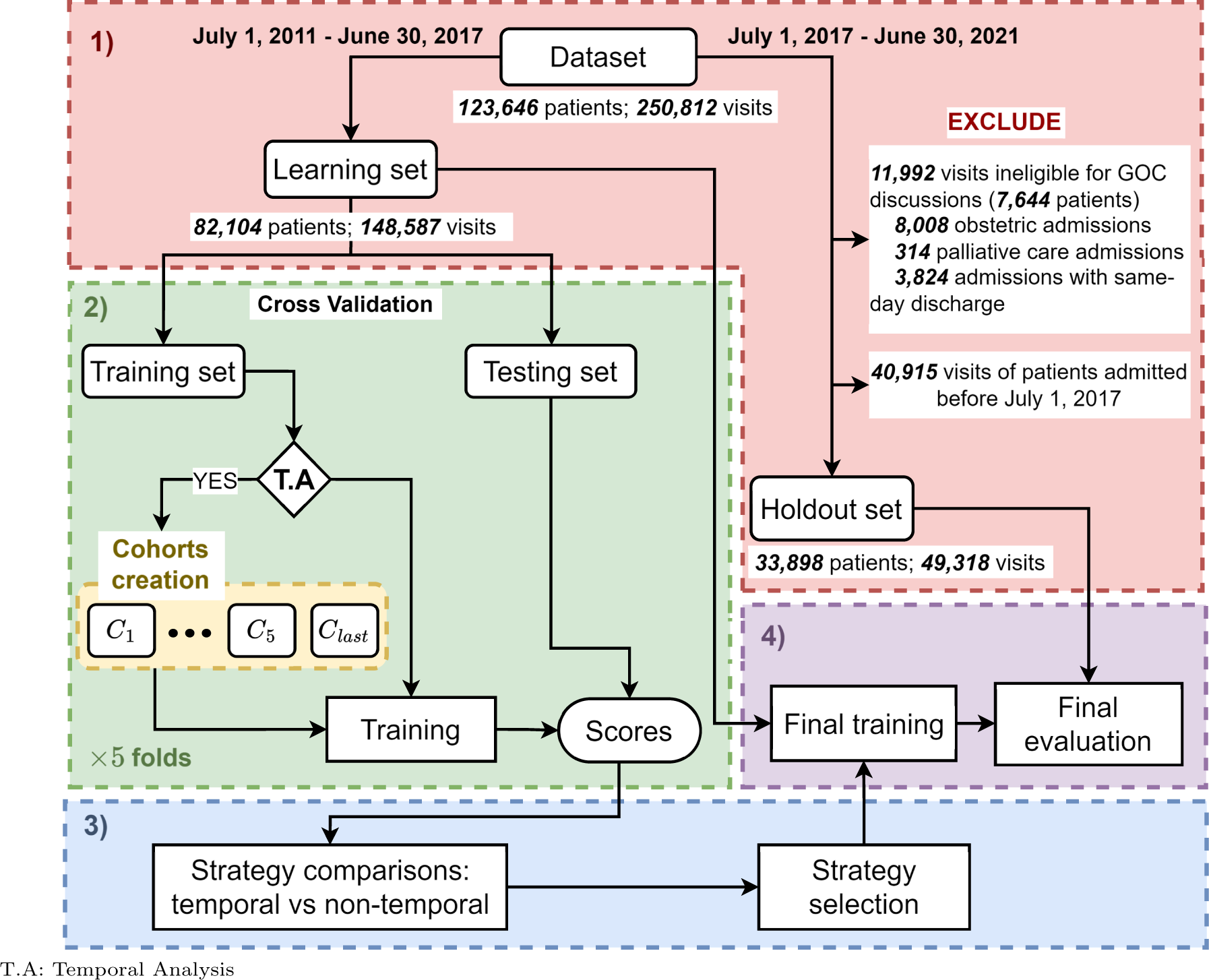
Experimental setup for model comparisons and final evaluation. 1) Temporal division of the dataset into a *learning set* and a *holdout set*. 2) Evaluation of the predictive performance of each model on 5 *testing sets* using a 5-fold cross-validation over the patients of the learning set. The same data splits are used for all models. Baseline models (RF, BLSTM) are trained on all the visits, while each temporal model LSTM_k_ comprising the ELSTM is trained using a temporal cohort *C*_*k*_. The scores are reported on specific patients of the *testing sets*. The training of each model includes the optimization of hyperparameters, except for the temporal models, for which we only optimize the hyperparameters of LSTM_last_. Details on hyperparameter optimization are shown in Appendix A. 3) Comparison of the temporal and non-temporal strategy and selection of the best strategy based on performance. The latter is measured using the mean and standard deviation of the scores on the 5 testing sets. 4) Final evaluation of the selected strategy on the holdout set. The final model predictions are then compared to usual care to quantify clinical utility

#### 2.3.3 Model selection procedure

In the model selection phase, we compare the performance of the ELSTM and baseline models to evaluate the benefits of incorporating the patients history in predicting their one-year mortality risk. To achieve this, we used a nested 5-fold cross-validation scheme. We partitioned the learning set using a 5-fold cross-validation into distinct training and test sets. Each of the training sets was subsequently separated into distinct inner training and inner test sets with an inner 5-fold cross-validation. The inner sets were entirely dedicated to optimize the hyperparameters of the models for each outer training fold. The data splitting was based on patients rather than visits, ensuring that each patient exclusively belonged to one set at a time.

To train the LSTM-based models, we created an additional validation set (as well as an inner validation set) for each of the 5 cross-validation splits, enabling us to track model performance through training epochs and proceed to early stopping if necessary. Each (inner) validation set was created by randomly sampling 10% of patients from the corresponding (inner) training set. At each (inner) training split, the baseline models were trained using all patients’ visits, while each LSTM_k_ part of the ELSTM was trained using a temporal cohort *C*_*k*_.

We assessed the benefits of the longitudinal data at each patient visit including their last visit available in our dataset, when we considered our patient’s medical trajectory completed. We define *V*_*t*_ as all the *t*^*th*^ visits of patients having at least *t* visits, and *V*_*t,last*_ as the last visits of patients having exactly *t* visits.

#### 2.3.4 Hyperparameters optimization

We optimized each model’s hyperparameters to find the best set leading to the highest scores. We trained each LSTM-based model using the Adam optimizer [22] with parameters *β*_1_ = 0.9 and *β*_2_ = 0.999, and a batch size of 100. We fixed the sizes of the fully connected layers to 2 and 1 respectively. Given that the ELSTM consists of multiple models, we chose to exclusively optimize the hyperparameters of LSTM_last_ and used the selected set to train each LSTM_k_. This way, we ensured consistent probability scales within the models constituting the ensemble model. For each optimized model, we sampled 100 sets of hyperparameters values from predefined search spaces, using a random sampler from the Optuna Python library [23]. Each set of hyperparameters values was evaluated by training the model with the 5 inner training sets and then measuring the AUROC on their respective inner testing sets. Here, the inner test sets included only the last visit of each patient. The set associated with the highest AUROC was selected to train the model on the whole training set of the outer loop. Models’ hyperparameters are provided in Appendix A.

#### 2.3.5 Final evaluation procedure

To evaluate the clinical utility of the best model selected in the previous phase, we compared its predictions to the usual care performed by clinicians on patients eligible for a GOC discussion. We aimed to quantify the gains and losses in terms of true positive and false positive alerts if this automated tool was used in a clinical decision support system to alert clinicians when a patient is identified as being at risk of one-year mortality. First, we extracted all CSOs of patients in the holdout set, and considered that a GOC occurred between a patient and a clinician (and that a patient at high risk of one-year mortality was identified by the clinical team) if a CSO was documented prior to the patient’s discharge, whether during the current admission or a previous one. Similarly to *Taseen and Ethier* [9], we defined:

- True Positives (TPs) as patients with a documented CSO who died within a year.
- False Positives (FPs) as patients with a documented CSO who survived beyond a year.
- False Negatives (FNs) as patients without a documented CSO who died within a year.
- True Negatives (TNs) as patients without a documented CSO who survived beyond a year.

Next, we trained the previously selected model using the entire learning set and compared its predictions, which would represent the actions suggested by the automated tool, to the usual care the patients from the holdout set received.

## 3 Results

The overall cohort consisted of 123,646 patients and 250,812 hospitalizations, with 15% of patients experiencing mortality within one year of their last admission. The learning set included 82,104 patients and 148,587 hospitalizations. For the holdout set, we excluded 40,915 hospitalizations belonging to previously admitted patients between 2011-2017, along with 7,644 patients and 11,992 hospitalizations ineligible for GOC discussions. Ultimately, the holdout set included 33,898 patients and 49,318 hospitalizations. Detailed descriptive analyses for each set can be found in Appendix B. Fig. 3a provides an overview of the proportion of mortality and survival per number of visits across the dataset. Patients who are frequently admitted to the hospital are generally fewer, but present a higher risk of one-year mortality. Fig. 3b shows the distribution of visits over time after the first hospitalization discharge. The second and third visits occur mainly in the first months following the first hospital discharge, while subsequent visits are increasingly scattered across time.

**Fig. 3.**
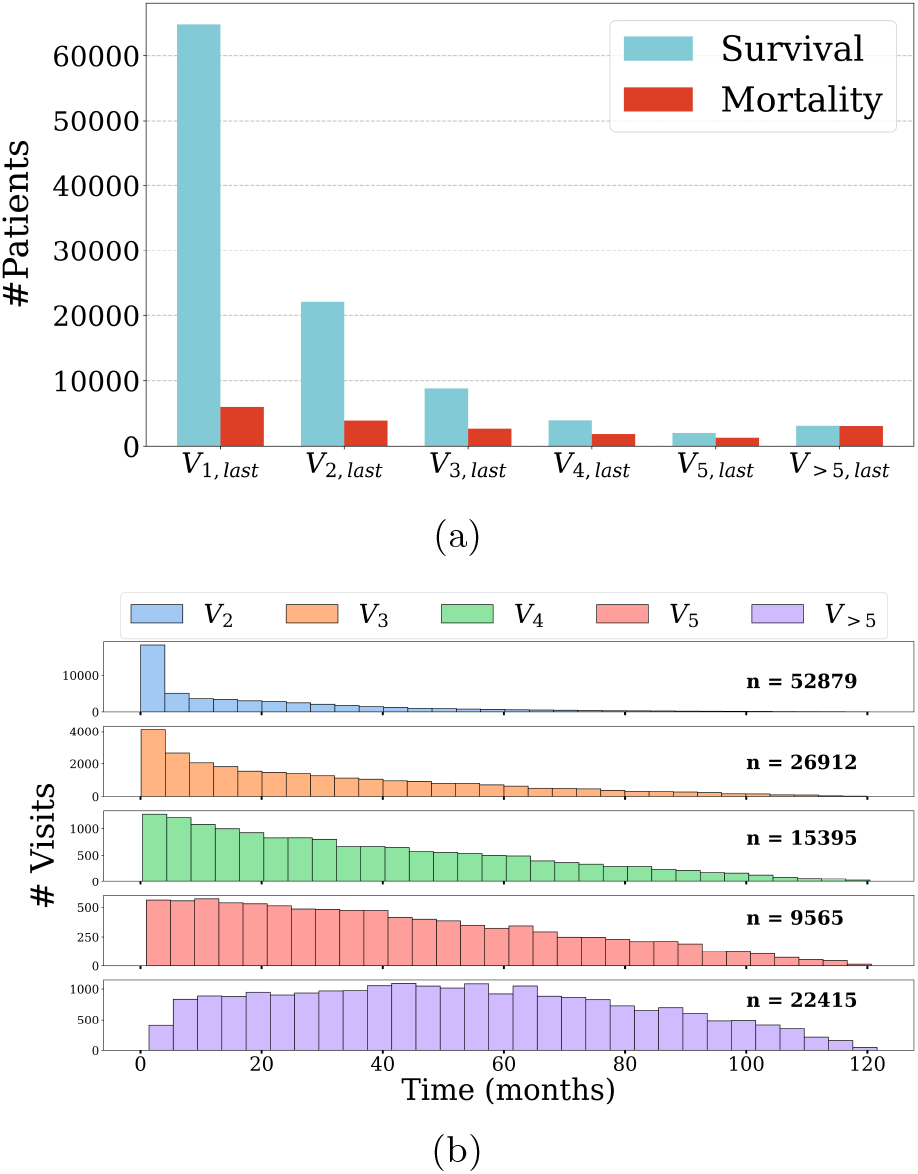
Distribution of visits across the entire dataset. (a) Proportions of survival and mortality per number of visits in the dataset. *V*_*t*,last_ represents all the patients with exactly *t* visits in the dataset and *V*_>*t*,last_ those with more than *t* visits. The number of patients decreases with the number of visits, in contrast to the mortality rate. (b) Distribution of visits over time after the first hospital discharge. *V*_*t*_ represents all the *t*^th^ visits in the dataset and *V*_>*t*_ all the visits after the *t*^th^ visit

### 3.1 Model selection on the learning set

In this part of our study, we explored the benefits of integrating patients’ historical data to predict their HOMR score. We assessed the baselines and the ELSTM on the learning set at various stages of patients’ hospital admissions, to understand the extent to which exploring patients’ history proves beneficial. As described earlier, we considered two groups of predictors: AdmDemo and AdmDemoDx.

Table 2a presents the performance of the baseline models and ELSTM for the last visit of each patient. The ELSTM outperforms the non-temporal models with a higher AUROC for all patient groups with both sets of predictors. Statistical tests revealed a significant overall improvement, except for *V*_5,last_ and *V*_>5,last_, where we note a higher variance due to fewer patients (∼ 300 and ∼ 500) that can diminish the statistical power of the test. Notably, even for patients without a historical record *V*_1,last_, the temporal model was effective – emphasizing that absence of recurrent visits serves as valuable insight. Experiments in Table 2b show that the impact of longitudinal data is less pronounced on intermediate visits, with non-statistically significant increases or decreases across multiple groups of patients, especially for AdmDemoDx predictors. Appendix C shows the performances of each individual LSTM_k_ within the ELSTM.

**Table 2:** Performance of the baseline models and the ELSTM on the testing sets of the learning set using both AdmDemo and AdmDemoDx predictors. RF: Random Forest; BLSTM: Basic LSTM; ELSTM: Ensemble LSTM. (a) Performance on the last visits of patients. (b) Performance on the last and intermediate visits of patients. Each testing set from the 5-fold cross-validation was divided into different groups of patients according to their number of visits to evaluate when temporal modeling is beneficial. Each group of patients included a patient at most once. The scores correspond to the *mean ± standard deviation* of the AUROC over the 5 testing sets. For each group of patients, the highest AUROC is highlighted in bold. Significant difference was quantified using the one-sided Wilcoxon signed-rank test [24]. Each *p*-value corresponds to the significance of improvement of the ELSTM over the best baseline model (RF or BLSTM) for a specific group of patients. *V*_*t*,last_ represents the *t*^th^ visits of patients having exactly *t* visits, *V*_>*t*,last_ the last visits of patients having more than *t* visits, *V*_*t*_ the *t*^th^ visits of patients having at least *t* visits, and *V*_>*t*_ one visit selected randomly that occurred after the *t*^th^ visit for patients having more than *t* visits.

| Patients group | AdmDemo |  |  | AdmDemoDx |  |  |
| --- | --- | --- | --- | --- | --- | --- |
|  | RF | BLSTM | ELSTM | RF | BLSTM | ELSTM |
| $V_{1,\text{last}}$ | $88.1 \pm 0.4$ | $88.2 \pm 0.3$ | **$88.7 \pm 0.3^{**}$** | $91.3 \pm 0.2$ | $91.4 \pm 0.2$ | **$91.8 \pm 0.3^{**}$** |
| $V_{2,\text{last}}$ | $88.1 \pm 0.5$ | $88.4 \pm 0.4$ | **$89.1 \pm 0.5^{**}$** | $92.1 \pm 0.6$ | $92.0 \pm 0.8$ | **$92.6 \pm 0.7^{**}$** |
| $V_{3,\text{last}}$ | $84.4 \pm 0.5$ | $85.2 \pm 0.5$ | **$86.8 \pm 0.9^{**}$** | $90.2 \pm 0.8$ | $90.4 \pm 1.1$ | **$91.1 \pm 1.0^{**}$** |
| $V_{4,\text{last}}$ | $80.3 \pm 1.1$ | $80.6 \pm 1.5$ | **$83.3 \pm 0.8^{**}$** | $87.5 \pm 0.9$ | $87.7 \pm 0.6$ | **$88.7 \pm 0.9^{**}$** |
| $V_{5,\text{last}}$ | $80.6 \pm 4.1$ | $81.7 \pm 3.9$ | **$82.5 \pm 4.4$** | $86.1 \pm 2.4$ | $85.8 \pm 2.3$ | **$87.0 \pm 2.6^*$** |
| $V_{>5,\text{last}}$ | $75.3 \pm 1.6$ | $75.0 \pm 1.4$ | **$75.6 \pm 1.4$** | $81.7 \pm 0.6$ | $81.8 \pm 0.5$ | **$82.4 \pm 0.6^*$** |
| Last visit<sup>a</sup> | $89.1 \pm 0.2$ | $89.2 \pm 0.3$ | **$89.8 \pm 0.2^{**}$** | $92.2 \pm 0.2$ | $92.3 \pm 0.2$ | **$92.6 \pm 0.3^{**}$** |

(b)
| Patients group | AdmDemo |  |  | AdmDemoDx |  |  |
| --- | --- | --- | --- | --- | --- | --- |
|  | RF | BLSTM | ELSTM | RF | BLSTM | ELSTM |
| $V_1$ | $84.2 \pm 0.4$ | **$84.5 \pm 0.3$** | $84.3 \pm 0.4$ | $88.2 \pm 0.2$ | **$88.4 \pm 0.2$** | $88.3 \pm 0.3$ |
| $V_2$ | $82.3 \pm 0.4$ | $82.6 \pm 0.2$ | **$82.8 \pm 0.4^{**}$** | **$87.2 \pm 0.5$** | $87.1 \pm 0.4$ | **$87.2 \pm 0.5$** |
| $V_3$ | $77.5 \pm 0.5$ | $78.0 \pm 0.6$ | **$79.3 \pm 0.7^{**}$** | $84.4 \pm 0.7$ | $84.6 \pm 0.8$ | **$84.7 \pm 0.7^{**}$** |
| $V_4$ | $74.5 \pm 0.5$ | $75.0 \pm 0.8$ | **$76.9 \pm 0.6^{**}$** | $81.9 \pm 0.5$ | $81.9 \pm 0.3$ | **$82.1 \pm 0.6$** |
| $V_5$ | $73.4 \pm 2.2$ | $73.7 \pm 2.1$ | **$74.3 \pm 1.7$** | $79.5 \pm 1.1$ | $79.4 \pm 1.4$ | **$79.9 \pm 1.3$** |
| $V_{>5}$ | $73.0 \pm 1.1$ | **$73.7 \pm 0.8$** | $72.9 \pm 0.7$ | **$79.9 \pm 0.8$** | $79.6 \pm 0.7$ | $79.1 \pm 1.6$ |
| Any visit<sup>b</sup> | $86.8 \pm 0.2$ | $87.0 \pm 0.2$ | **$87.3 \pm 0.3^{**}$** | $90.3 \pm 0.1$ | $90.3 \pm 0.2$ | **$90.6 \pm 0.2^{**}$** |

<sup>a</sup>Last visit: last visits of all patients.
<sup>b</sup>Any visit: one visit per patient in the testing set selected randomly.
\* *p*-value < 0.1.
\*\* *p*-value < 0.05.

Next, the group of predictors with fewer variables (AdmDemo) achieved acceptable results for all patient sets with a predictably lower AUROC compared to AdmDemoDx (Tables 2a and 2b). The AdmDemo feature set seems to benefit more from the longitudinal data, as we observed a higher AUROC improvement across all patient sets compared to the AdmDemoDx feature set when using the ELSTM. This emphasizes the importance of incorporating longitudinal data in cases where variables about the patient’s precondition are not available (e.g., comorbidity diagnoses), and the ability of such longitudinal data to provide a more comprehensive understanding of the patients through their history.

In addition, the ELSTM revealed comparable AUROCs on patients admitted later in time to the AUROCs observed in the learning set (see Appendix D), thereby demonstrating its temporal validity. Overall, the ELSTM achieved the best performance for most patient groups, particularly on their last visits completing their medical trajectory. These results highlight the gains from integrating longitudinal patient data to predict the HOMR score.

### 3.2 Final evaluation on patients eligible for a GOC discussion

In this section, we compared the ELSTM using AdmDemo or AdmDemoDx predictors with the usual care performed by clinicians for each patient in the holdout set. We optimized the decision threshold for considering a patient at risk of one-year mortality by maximizing the Youden’s *J*-index [25]. We set it at 0.34 for ELSTM-AdmDemo and 0.17 for ELSTM-AdmDemoDx.

Results in Table 3 revealed that the ELSTM with AdmDemo predictors constitutes an automated tool with similar predictions to the usual care performed by clinicians, with overall good precision and a low rate of inappropriate alerts relative to daily clinical practice. We also observe that, although the ELSTM with AdmDemoDx predictors achieved the highest AUROC, the model is less sensitive and detects slightly fewer patients who actually died within a year of their admission (Fig. 4a). Nevertheless, this model considerably reduced the number of false positive notifications and increased the precision. The calibration curves in Fig. 4b support this result, by showing a tendency of ELSTM-AdmDemo to overestimate the risk of death compared to ELSTM-AdmDemoDx.

**Table 3:** Comparisons of the final ELSTM with AdmDemo and AdmDemoDx predictors to the usual care performed by clinicians on patients of the holdout set. The scores correspond to the *mean* ± *standard deviation* of the metric over 100 bootstraps drawn with replacement. The highest value for each metric is highlighted in bold.

|  | Any visit<sup>a</sup> |  |  | Last visit<sup>b</sup> |  |  |
| --- | --- | --- | --- | --- | --- | --- |
|  | Clinicians | AdmDemo | AdmDemoDx | Clinicians | AdmDemo | AdmDemoDx |
| AUROC | *N.A* | 83.0 ± 0.3 | **87.2 ± 0.3** | *N.A* | 85.3 ± 0.3 | **89.0 ± 0.3** |
| Sensitivity | 74.9 ± 0.6 | **75.2 ± 0.7** | 73.1 ± 0.7 | **81.7 ± 0.5** | 80.5 ± 0.6 | 77.9 ± 0.6 |
| Specificity | 72.2 ± 0.2 | 76.0 ± 0.2 | **84.1 ± 0.2** | 70.4 ± 0.3 | 75.1 ± 0.2 | **83.6 ± 0.2** |
| Precision | 24.8 ± 0.4 | 27.7 ± 0.4 | **36.0 ± 0.5** | 28.0 ± 0.4 | 31.3 ± 0.4 | **40.1 ± 0.5** |
| NPV | 95.9 ± 0.1 | **96.2 ± 0.1** | **96.2 ± 0.1** | **96.5 ± 0.1** | **96.5 ± 0.1** | 96.4 ± 0.1 |

<sup>a</sup>Any visit: one visit per patient in the holdout set selected randomly.
<sup>b</sup>Last visit: last visits of all patients.

**Fig. 4.**
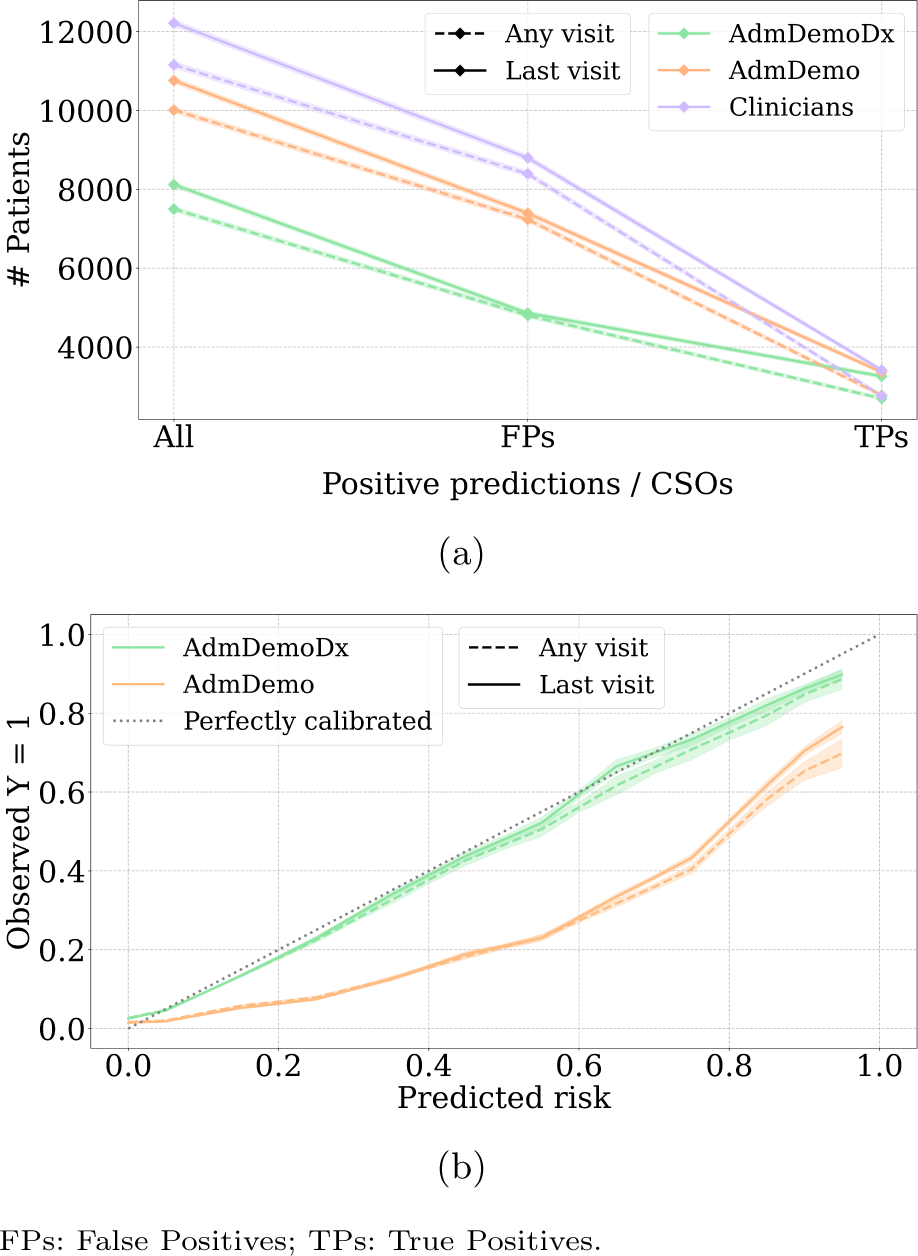
Analyses of the final ELSTM tested on the holdout set with both AdmDemo and AdmDemoDx predictors. Shaded regions indicate variations within one standard deviation of the mean over 100 bootstraps. (a) Number of positive predictions by the ELSTM with AdmDemo and AdmDemoDx predictors, and CSOs documented by clinicians. The ELSTM shows a reduced number of false positives and a slight loss in the number of true positives. (b) ELSTM calibration curves with AdmDemo and AdmDemoDx predictors. We used interpolation to unify the predicted risk bins over the 100 bootstraps and generate a mean calibration curve with its variations. We show in Appendix C the 100 calibration curves for each ELSTM in the bootstrap sampling. The ELSTM-AdmDemoDx is almost identical to a perfectly calibrated model, while the ELSTM-AdmDemo tends to overestimate the risk of mortality

Finally, we analyzed the evolution of importance for each group of features along with the number of visits per patient in the ELSTM-AdmDemoDx. Post-hoc analyses of the importance assigned to each feature by a model provided important insights into their impact on the predicted scores. Fig. 5 illustrates that as the number of patient visits increases, the importance of longitudinal information grows, and the model relies on predictors from both current and previous admissions to predict mortality risk. Conversely, as the number of visits decreases, the model relies more heavily on demographic information to make its predictions. This highlights the importance of using longitudinal data for patients with a long medical history, and is consistent with clinical reality, where the frequently admitted patients’ prognoses have a higher dependence on their overall health history than on their demographics. Feature importance and the overall performance of the ELSTM did not vary when we included the time gap between current and previous admissions (see Appendix E), demonstrating that the model was able to learn this information solely through the content of longitudinal records.

**Fig. 5.**
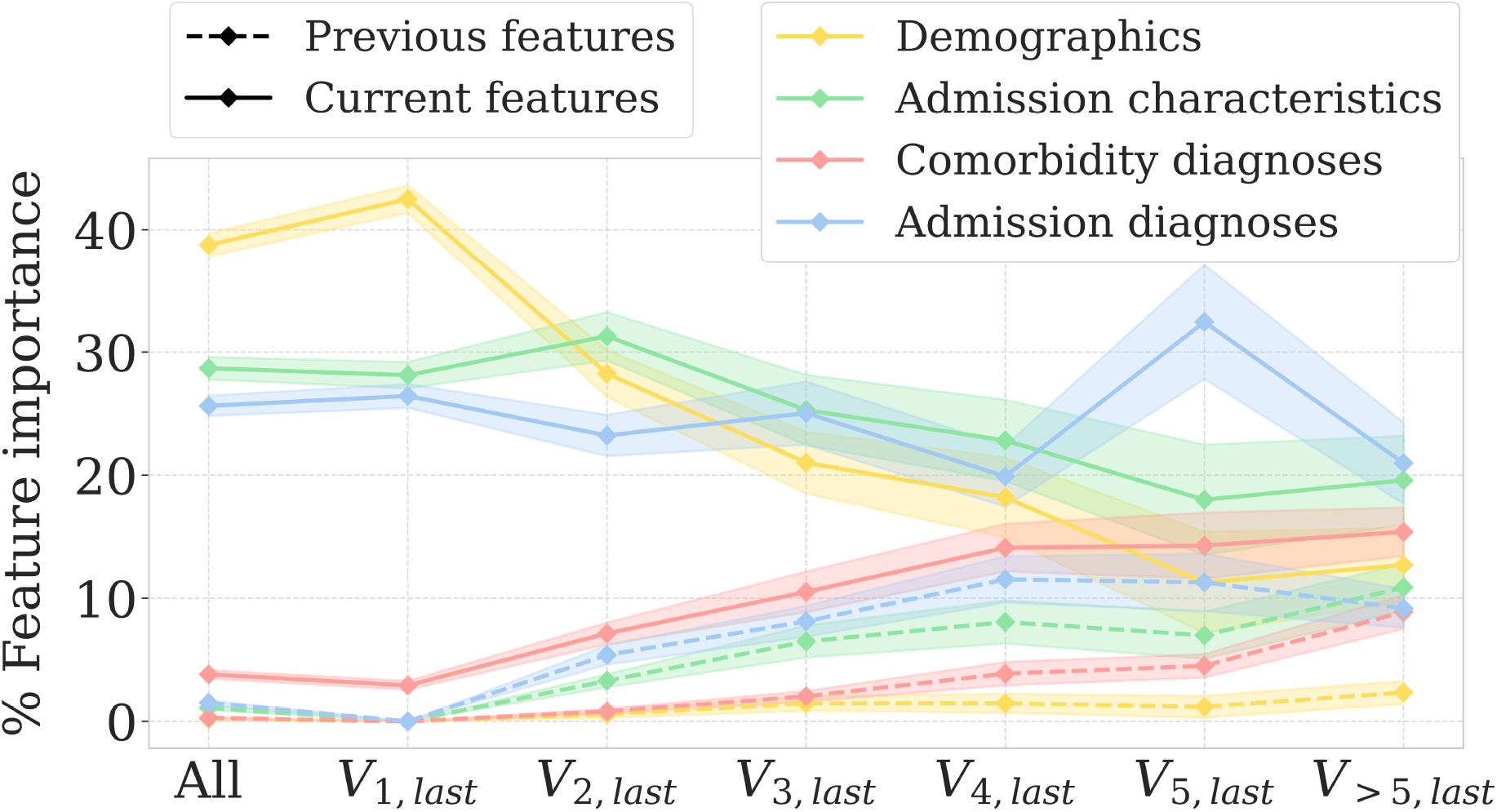
Post-hoc analyses of feature importance of the final ELSTM trained with AdmDemoDx predictors. Importance of each feature is computed using feature permutation [26] over 100 bootstraps. Shaded regions indicate variations within one standard deviation of the mean over 100 bootstraps. Importance of previous features increases as the size of patients’ history gets longer

## 4 Discussion

Recent years have seen efforts dedicated to developing automated models identifying patients at high risk of mortality, in order to improve end-of-life care and align patient preferences with the provided care. Recent works have explored the use of machine learning models to integrate patients’ longitudinal data in several clinical contexts [27–29], and presented interesting improvements over single-visit models. However, to date, these techniques have not been used for models predicting the HOMR score to enhance palliative care. This study introduces the Ensemble Long Short-Term Memory (ELSTM), a recurrent neural network-based ensemble model that integrates both admission and historical patient data to automatically identify individuals at an elevated risk of one-year mortality. The aim is to prompt the clinical team for end-of-life interventions, such as GOC discussions.

Firstly, we developed the ELSTM, an ensemble model built upon the LSTM neural network, that leverages information learned by different LSTMs at various stages of a patient’s admission. We applied the ELSTM to patients with varying numbers of visits and estimated their HOMR score. We used patient self-reported predictors available upon admission (AdmDemo), as well as other comorbidity diagnoses available in patients’ EHR and admission diagnoses documented later during their stay (AdmDemoDx). A significant improvement in AUROC, the standard evaluation metric in the literature for measuring the discriminative power of the HOMR score [13], was observed across the majority of patients groups using both sets of predictors. Within the LSTM-based neural network, we believe the longitudinal data contributed to mortality prediction in two aspects. First, frequent visits to the hospital (i.e., more longitudinal data) likely indicate an increasing severity of illness, thus a higher risk of death. Second, the characteristics of each previous hospitalization (i.e., the content of longitudinal data) provide a superior holistic view of the patient’s overall condition. Thus, the importance of previous data grows with the length of a patient’s history. These two aspects allow each LSTM to learn long- and short-term longitudinal patterns to accurately identify patients at high risk of one-year mortality.

Next, we compared the ELSTM to the usual care provided by clinicians to the population of interest eligible for end-of-life interventions. Both AdmDemo and AdmDemoDx strategies revealed considerable benefits as an automated alert prevalence tool of patients at high risk of one-year mortality in a clinical decision support system. More specifically, the ELSTM using AdmDemo predictors facilitates real-time data acquisition, as it requires fewer variables, all available immediately upon admission and can be self-reported. In addition, the model revealed similar results to human decision-making, and is hence useful in hospitals where diagnoses are encoded post-discharge as in previous studies [15, 16]. On the other hand, even though the ELSTM using AdmDemoDx predictors is more challenging in terms of data acquisition, it can significantly reduce false positive notifications and therefore the risk of an alert fatigue, making it a suitable candidate for deployment in a clinical decision support system.

We have identified several limitations worth addressing in future studies. Firstly, while the overall AUROC of the ELSTM with the AdmDemoDx feature set ranges between 0.87-0.89, an examination of population subgroups shows that the oldest patients, and potentially those with the most complicated medical conditions, are less accurately predicted (see Appendix E). To better identify patients at high risk of mortality, models used on these patients should include not only administrative and diagnostic variables routinely collected on admission, but also admission-specific clinical variables such as vital signs, laboratory and imaging tests. Secondly, our evaluation of clinical utility assumes that a clinician would engage in a GOC discussion and document a CSO for all and only those patients suspected to be at high risk of death. However, this assumption has limitations. Not all high-risk patients may have the opportunity for a GOC discussion due to a lack of resources or time. Additionally, clinicians may document a patient’s CSO not only based on their risk of mortality but also on the potential need for escalated care requiring intubation or ventilation. Thirdly, although the AUROC is the main metric in the literature to evaluate models predicting the HOMR score, model selection in clinical settings should primarily maximize clinical utility, which is extremely context-dependent (based on individual hospital services and resources, typical patient origin and profile, severity of admissions, length of stay, etc). Fourthly, although our model demonstrated an acceptable temporal validity, it was not validated using external datasets. Let us however note that the HOMR prediction score has been externally validated in a previous study [14]. Finally, it is important to acknowledge that predicting a patient at high risk of mortality does not guarantee an effective GOC discussion. Future research should therefore investigate the actual impact of early detection of these patients on the quality of their end-of-life care.

## 5 Conclusion

In this work, we developed an ELSTM, an ensemble recurrent neural network-based approach leveraging information available across different patient hospitalizations. We evaluated our model using data collected routinely during hospital admissions to predict the Hospital One-year Mortality Risk (HOMR) score and to identify individuals who might benefit from end-of-life discussions with healthcare providers. Our model outperformed existing approaches both when using only admission demographics and administrative variables as predictors (AdmDemo), and when integrating diagnoses as well (AdmDemoDx). Our study highlights the rich data potential available in patients’ medical records, emphasizing their ability to generate predictive models for enhancing patient care, throughout the life spectrum and at the end of life.

## Data Availability

Software code allowing to run the experiments used
to produce the results presented in this work is freely shared under the GNU General Public License v3.0 on the GitHub website at: https://github.com/MEDomics-UdeS/POYM. The hospitalization data analysed during the current study are not publicly available for confidentiality purposes overseen by the IRB (Institutional Review Board of the CIUSSS de l Estrie - CHUS Nagano \#2022-4409). However, a randomly generated dataset with the same format as used in our experiments is publicly shared in our GitHub repository to test the code implemented for this work.

## Declarations

### Ethics approval and consent to participate

Institutional Review Board approval was obtained prior to data acquisition (Institutional Review Board of the CIUSSS de l’Estrie—CHUS Nagano #2022-4409). Given the retrospective nature of the study, the Director of Professional Services of CIUSSS de l’Estrie-CHUS waived the requirement for signed consent from participants.

### Consent for publication

All authors had full access to all the data in the study and accept responsibility to submit for publication.

### Data and code availability

Software code allowing to run the experiments used to produce the results presented in this work is freely shared under the GNU General Public License v3.0 on the GitHub website at: https://github.com/MEDomics-UdeS/POYM. The hospitalization data analysed during the current study are not publicly available for confidentiality purposes overseen by the IRB (Institutional Review Board of the CIUSSS de l’Estrie—CHUS Nagano #2022-4409). However, a synthetic dataset generated using the AVATAR method [30] in partnership with Octopize^2^ is publicly shared on the Zenodo website at: https://zenodo.org/doi/10.5281/zenodo.12954672. The publication of this synthetic dataset has been approved under project #2022-4409 with form #F2H-60510.

### Authors’ contributions

Authors’ initials are used to designate them.

Conceptualization: HL, MV

Data curation: HL, RT

Formal Analysis: HL

Funding acquisition: MV

Investigation: HL, MV, RT

Methodology: HL, MV

Project administration: HL, MV

Resources: MV

Software: HL, NR

Supervision: MV, DP

Validation: HL, DP

Visualization: HL

Writing – original draft: HL

Writing – review & editing: HL, MV, DP, RT, NR

## Acknowledgements

We thank Jean-François Ethier, Associate Professor in the Department of Medicine at the Université de Sherbrooke, for data collection. We also thank Olivier Lefebvre, PhD student at Université de Sherbrooke, and Mahdi Ait Lhaj Loutfi, Master’s student at Université de Sherbrooke, for helpful comments and suggestions throughout the project.

## Appendix A. Hyperparameters

This section provides further information on the hyperparameter optimization process (Fig. 6), including the hyperparameters of the models and their respective search spaces (Table 4 and 5).

**Table 4:** Random Forest’ hyper-parameters. The hyperparameters that are not mentioned were set as the default ones from the *scikitlearn* wrapper interface of version 0.8.0 of the *skranger* library^3^. The *weight* hyperparameter represents the weight of the positive class.

| Hyperparameter | Search space |
| --- | --- |
| n_estimators | {128, 256, ..1024} |
| mtry | {10, 15, 20} |
| min_node_size | {10, 20, ..80} |
| weight | [0.1, 0.9] |

**Table 5:**
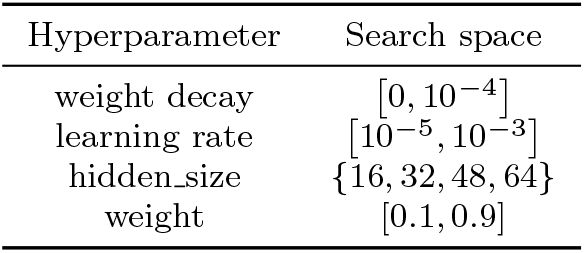
Hyperparameters of LSTM-based models (BLSTM and LSTM_k_). The *weight decay* refers to the coefficient multiplying the ℒ_2_ penalty in the cross entropy loss. The *learning rate* refers to the initial learning rate given to the Adam optimizer [22] at the beginning of the training. The *hidden size* refers to the number of neurons in the hidden layer. The *weight* hyperparameter represents the weight of the positive class.

| Hyperparameter | Search space |
| --- | --- |
| weight decay | $[0, 10^{-4}]$ |
| learning rate | $[10^{-5}, 10^{-3}]$ |
| hidden_size | $\{16, 32, 48, 64\}$ |
| weight | $[0.1, 0.9]$ |

**Fig. 6.**
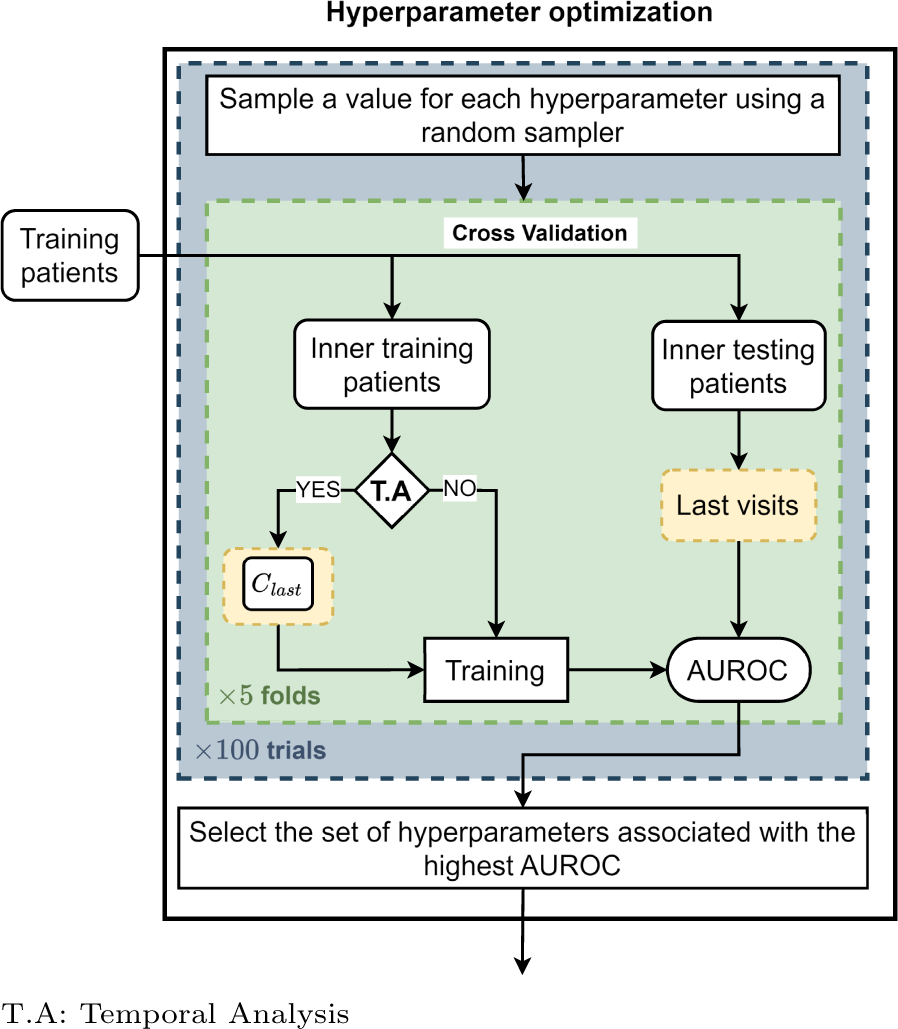
Hyperparameter optimization for each model. The process is performed automatically using a random sampler from predefined search spaces for each hyperparameter, within the framework of the Optuna [23] Python library. The training patients are divided using a 5-fold cross-validation into inner training patients and inner testing patients. Temporal models are trained using the temporal cohort *C*_last_. All models are tested on the last visits of patients. A total of 100 sets of hyperparameter values are sampled sequentially and evaluated on the same inner testing patients. The performance is measured using the mean of the AUROC on the 5 inner testing sets. The set of hyperparameters associated with the highest AUROC is used to train the model on the outer training patients

## Appendix B. Descriptive analyses

This section provides detailed descriptive analyses of the demographic and admission characteristic features, as well as four major comorbidities, across the full dataset (Table 6), the learning set (Table 7), and the holdout set (Table 8). We present the mode of each categorical feature along with its proportion in the dataset, and the mean of each continuous feature along with its standard deviation. The *p*-values are computed using the Welch’s t-test [31] for continuous features (age, ambulance admission count, ED visit count, and weeks recently hospitalized) and the Pearson’s chi-squared test [32] for categorical and binary features, using the *scipy* [33] Python library.

**Table 6:** Descriptive analysis of the demographics and admission characteristics features along with four major comorbidities on the full dataset.

| Variable | All<br>(n=250,812) | Survivors<br>(n=214,095) | Deceased<br>(n=36,717) | *p*-value |
| --- | --- | --- | --- | --- |
| Demographics |  |  |  |  |
| Age | 61.12 ± 20.07 | 58.98 ± 20.18 | 73.62 ± 13.93 | < 0.001 |
| Sex | Female (54 %) | Female (55 %) | Male (54 %) | < 0.001 |
| Admission characteristics |  |  |  |  |
| Ambulance admission | 0 (71 %) | 0 (74 %) | 1 (51 %) | < 0.001 |
| Flu season | 0 (75 %) | 0 (75 %) | 0 (74 %) | < 0.001 |
| ICU admission | 0 (97 %) | 0 (97 %) | 0 (95 %) | < 0.001 |
| Urgent 30-d readmission | 0 (90 %) | 0 (92 %) | 0 (79 %) | < 0.001 |
| Ambulance admissions count | 0.23 ± 0.75 | 0.17 ± 0.64 | 0.56 ± 1.19 | < 0.001 |
| ED visits count | 0.8 ± 1.52 | 0.7 ± 1.42 | 1.38 ± 1.89 | < 0.001 |
| Weeks recently hospitalized | 0.29 ± 0.99 | 0.21 ± 0.85 | 0.71 ± 1.5 | < 0.001 |
| Living status | Home (48 %) | Unknown (50 %) | Home (59 %) | < 0.001 |
| Admission service | Cardiology (13 %) | Obstetrics (15 %) | I.M<sup>a</sup> (14 %) | < 0.001 |
| Admission type | Urgent (65 %) | Urgent (60 %) | Urgent (89 %) | < 0.001 |
| Major comorbidities |  |  |  |  |
| Dementia | 0 (97 %) | 0 (98 %) | 0 (93 %) | < 0.001 |
| Congestive heart failure | 0 (94 %) | 0 (95 %) | 0 (86 %) | < 0.001 |
| Metastatic solid cancer | 0 (98 %) | 0 (99 %) | 0 (91 %) | < 0.001 |
| Asthma | 0 (97 %) | 0 (97 %) | 0 (96 %) | < 0.001 |

<sup>a</sup>I.M: Internal Medicine.

**Table 7:** Descriptive analysis of the demographics and admission characteristics features along with four major comorbidities on the learning set.

| Variable | All<br>(n=148,587) | Survivors<br>(n=127,996) | Deceased<br>(n=20,591) | *p*-value |
| --- | --- | --- | --- | --- |
| Demographics |  |  |  |  |
| Age | 60.52 ± 20.27 | 58.45 ± 20.36 | 73.34 ± 13.99 | < 0.001 |
| Sex | Female (54 %) | Female (56 %) | Male (54 %) | < 0.001 |
| Admission characteristics |  |  |  |  |
| Ambulance admission | 0 (70 %) | 0 (74 %) | 1 (52 %) | < 0.001 |
| Flu season | 0 (75 %) | 0 (75 %) | 0 (74 %) | 0.003 |
| ICU admission | 0 (97 %) | 0 (97 %) | 0 (95 %) | < 0.001 |
| Urgent 30-d readmission | 0 (91 %) | 0 (92 %) | 0 (79 %) | < 0.001 |
| Ambulance admissions count | 0.24 ± 0.78 | 0.18 ± 0.66 | 0.59 ± 1.26 | < 0.001 |
| ED visits count | 0.86 ± 1.57 | 0.75 ± 1.47 | 1.52 ± 1.99 | < 0.001 |
| Weeks recently hospitalized | 0.29 ± 0.98 | 0.22 ± 0.86 | 0.73 ± 1.47 | < 0.001 |
| Living status | Unknown (49 %) | Unknown (52 %) | Home (61 %) | < 0.001 |
| Admission service | Obstetrics (13 %) | Obstetrics (15 %) | F.M<sup>a</sup> (16 %) | < 0.001 |
| Admission type | Urgent (64 %) | Urgent (60 %) | Urgent (89 %) | < 0.001 |
| Major comorbidities |  |  |  |  |
| Dementia | 0 (97 %) | 0 (98 %) | 0 (93 %) | < 0.001 |
| Congestive heart failure | 0 (94 %) | 0 (95 %) | 0 (85 %) | < 0.001 |
| Metastatic solid cancer | 0 (98 %) | 0 (99 %) | 0 (91 %) | < 0.001 |
| Asthma | 0 (97 %) | 0 (97 %) | 0 (96 %) | < 0.001 |

<sup>a</sup>F.M: Family Medicine.

**Table 8:**
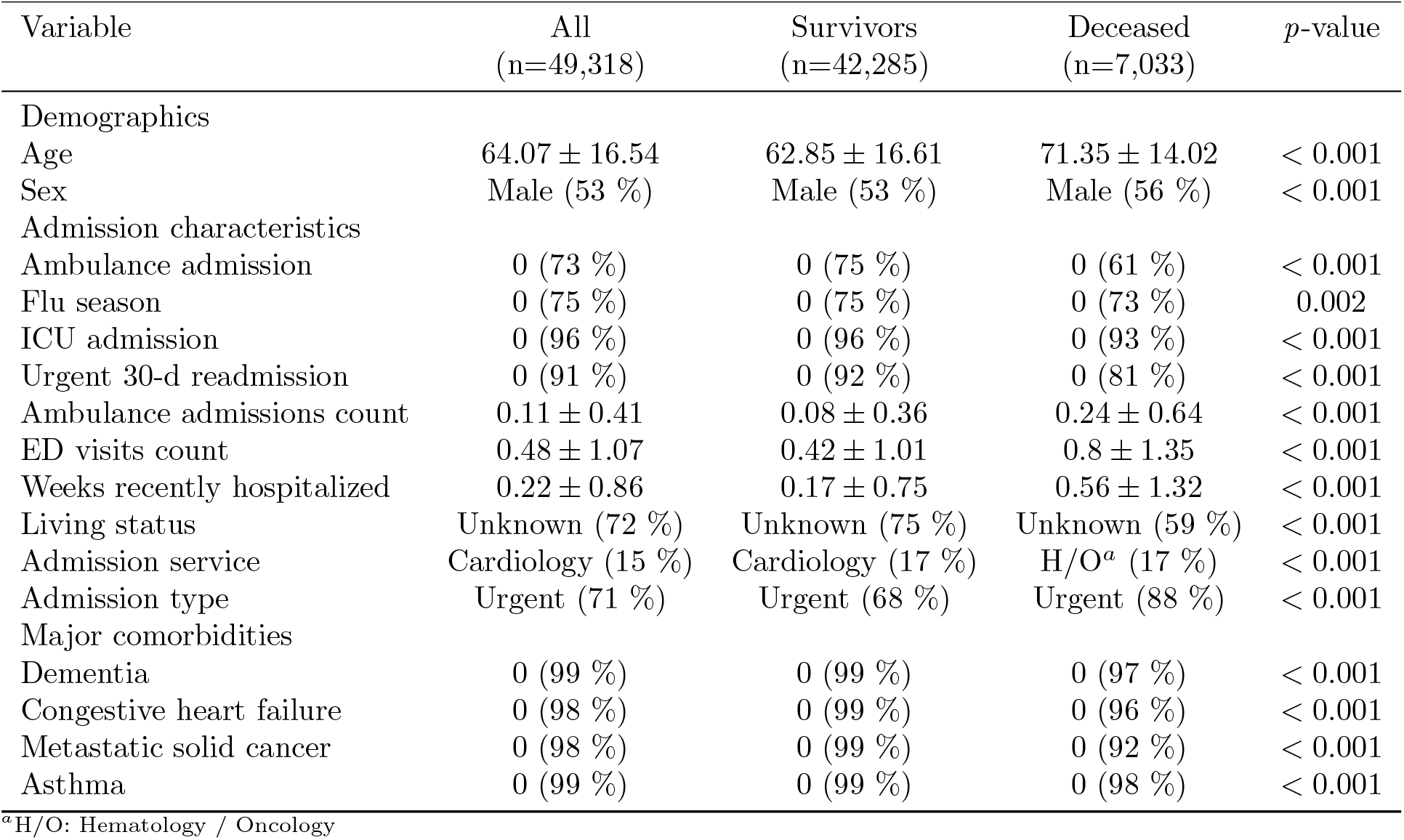
Descriptive analysis of the demographics and admission characteristics features along with four major comorbidities on the holdout set.

## Appendix C. Detailed performance of the ELSTM

We present the performance of each LSTM_k_ constituting the ELSTM on patients of the learning set in Fig. 7.

**Fig. 7.**
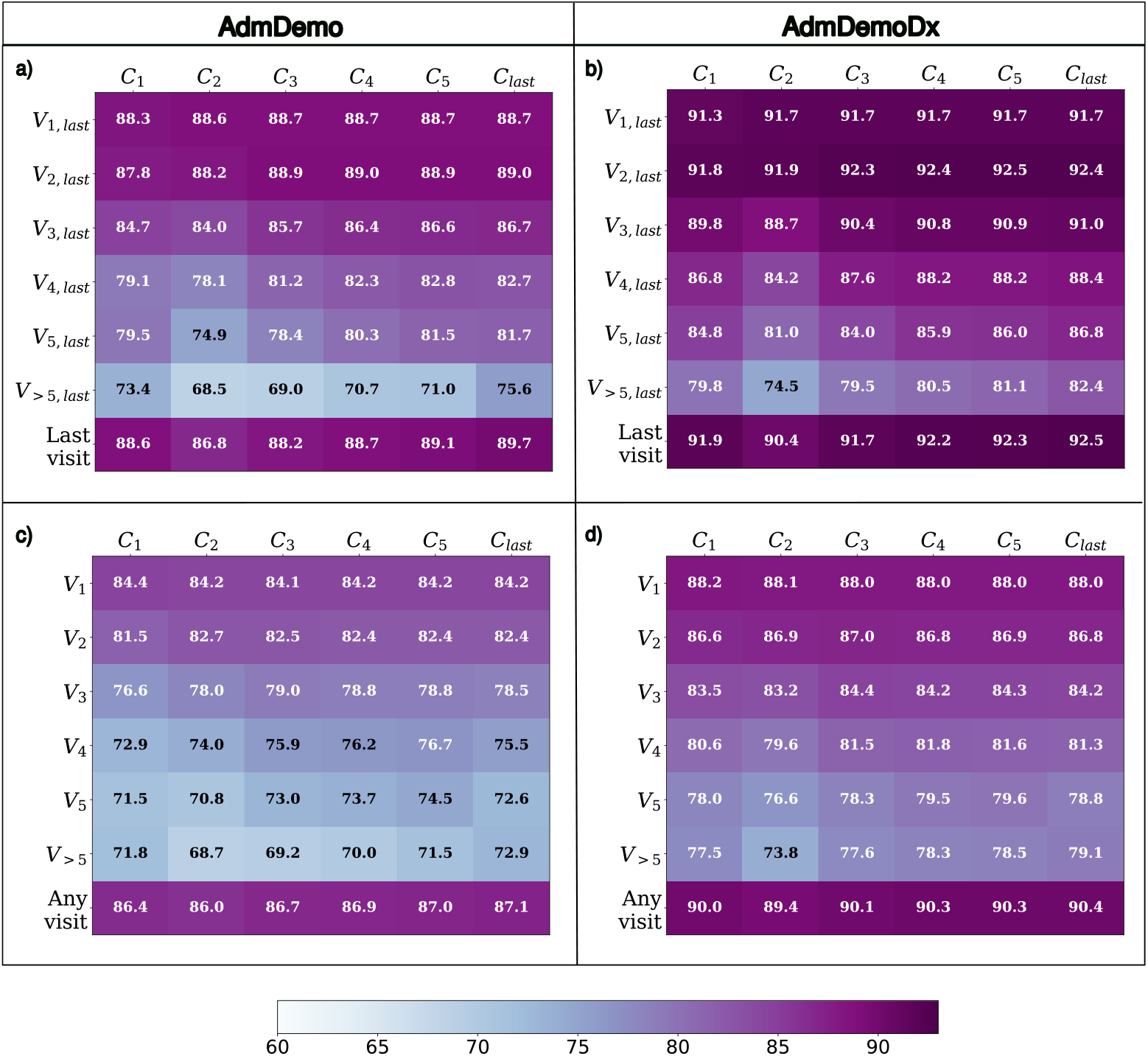
Performance of each LSTM_k_ trained with a cohort *C*_*k*_ on different groups of patients. (a) and (b) Performance on the last visits of patients. (c) and (d) Performance on the last and intermediate visits of patients. The rows represent the testing patients and the columns represent the training cohorts. The scores in the intersection of a row and a column correspond to the *mean* of the AUROC over the 5 folds of cross-validation of an LSTM trained with the corresponding cohort and tested on the corresponding patients

Additionally, we provide the calibration curves of the final ELSTM, tested on 100 bootstraps of the holdout set, at each bootstrap in Fig. 8.

**Fig. 8.**
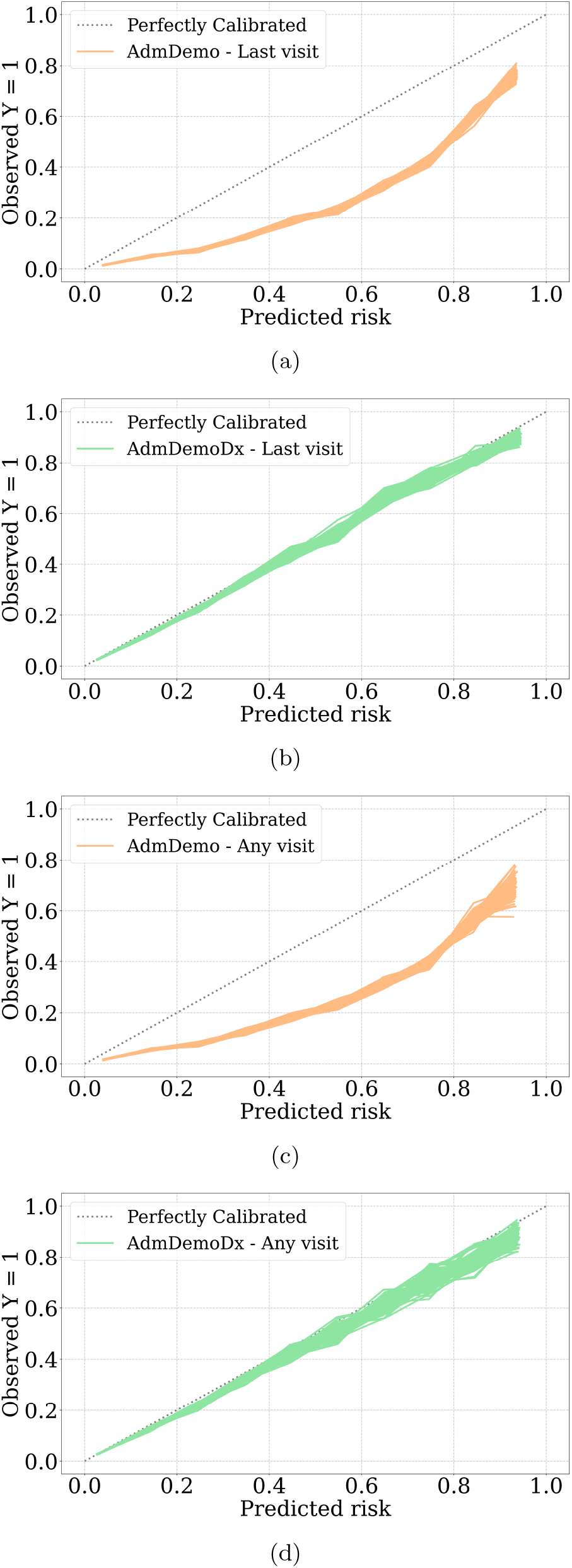
Calibration curves of the ELSTM with AdmDemo and AdmDemoDx predictors for each of the 100 bootstraps on the holdout set

## Appendix D. Temporal validity

We train the ELSTM with patients admitted between July 1, 2011 and June 30, 2017 and tested it on patients who are only admitted between July 1, 2017 and June 30, 2021, without excluding those ineligible for a GOC discussion. The testing set included 41,542 patients and 61,310 hospitalizations. The goal is to evaluate the ELSTM when using the same rules for excluding visits for training and testing, but with data from different time periods. Results in Table 9 reveal an acceptable temporal validity, with AUROCs comparable to those observed in the learning set (Table 2).

**Table 9:** Temporal validity of the ELSTM without excluding patients not eligible for GOC discussions. (a) Performance on the last visits of patients. (b) Performance on the last and intermediate visits of patients. The scores correspond to the *mean* ± *standard deviation* of the AUROC over 100 bootstraps.

| (a) |  |  | (b) |  |  |
| --- | --- | --- | --- | --- | --- |
| Patients group | AdmDemo | AdmDemoDx | Patients group | AdmDemo | AdmDemoDx |
| $V_{1,last}$ | 86.3 ± 0.4 | 89.6 ± 0.3 | $V_1$ | 84.0 ± 0.3 | 87.8 ± 0.3 |
| $V_{2,last}$ | 88.3 ± 0.5 | 91.6 ± 0.4 | $V_2$ | 84.4 ± 0.4 | 88.1 ± 0.4 |
| $V_{3,last}$ | 85.8 ± 0.9 | 88.7 ± 0.9 | $V_3$ | 81.5 ± 0.8 | 85.6 ± 0.7 |
| $V_{4,last}$ | 84.0 ± 1.4 | 88.9 ± 1.1 | $V_4$ | 79.0 ± 1.0 | 84.7 ± 1.0 |
| $V_{5,last}$ | 86.1 ± 1.8 | 90.4 ± 1.6 | $V_5$ | 78.8 ± 1.5 | 84.3 ± 1.4 |
| $V_{>5,last}$ | 81.5 ± 1.7 | 85.4 ± 1.5 | $V_{>5}$ | 79.0 ± 1.8 | 83.4 ± 1.5 |
| Last visit | 88.2 ± 0.2 | 91.1 ± 0.2 | Any visit | 86.3 ± 0.3 | 89.5 ± 0.2 |

## Appendix E. Additional experimental results

We trained the ELSTM model with the time gap between current and previous admissions included as an additional predictor to assess its impact on predicting patient mortality risk. The overall performance (Table 10) and feature importance (Fig. 9) remained consistent with the results obtained using the original set of predictors.

**Table 10:** Performance of the ELSTM with AdmDemoDx predictors when including the time gap between current and previous admissions as a predictor. The scores correspond to the *mean* ± *standard deviation* of the metric over 100 bootstraps.

|  | Any visit | Last visit |
| --- | --- | --- |
| AUROC | $87.1 \pm 0.3$ | $88.9 \pm 0.3$ |
| Sensitivity | $75.3 \pm 0.6$ | $79.5 \pm 0.6$ |
| Specificity | $82.2 \pm 0.2$ | $82.0 \pm 0.2$ |
| Precision | $34.1 \pm 0.5$ | $38.4 \pm 0.5$ |
| NPV | $96.5 \pm 0.1$ | $96.6 \pm 0.1$ |

**Fig. 9.**
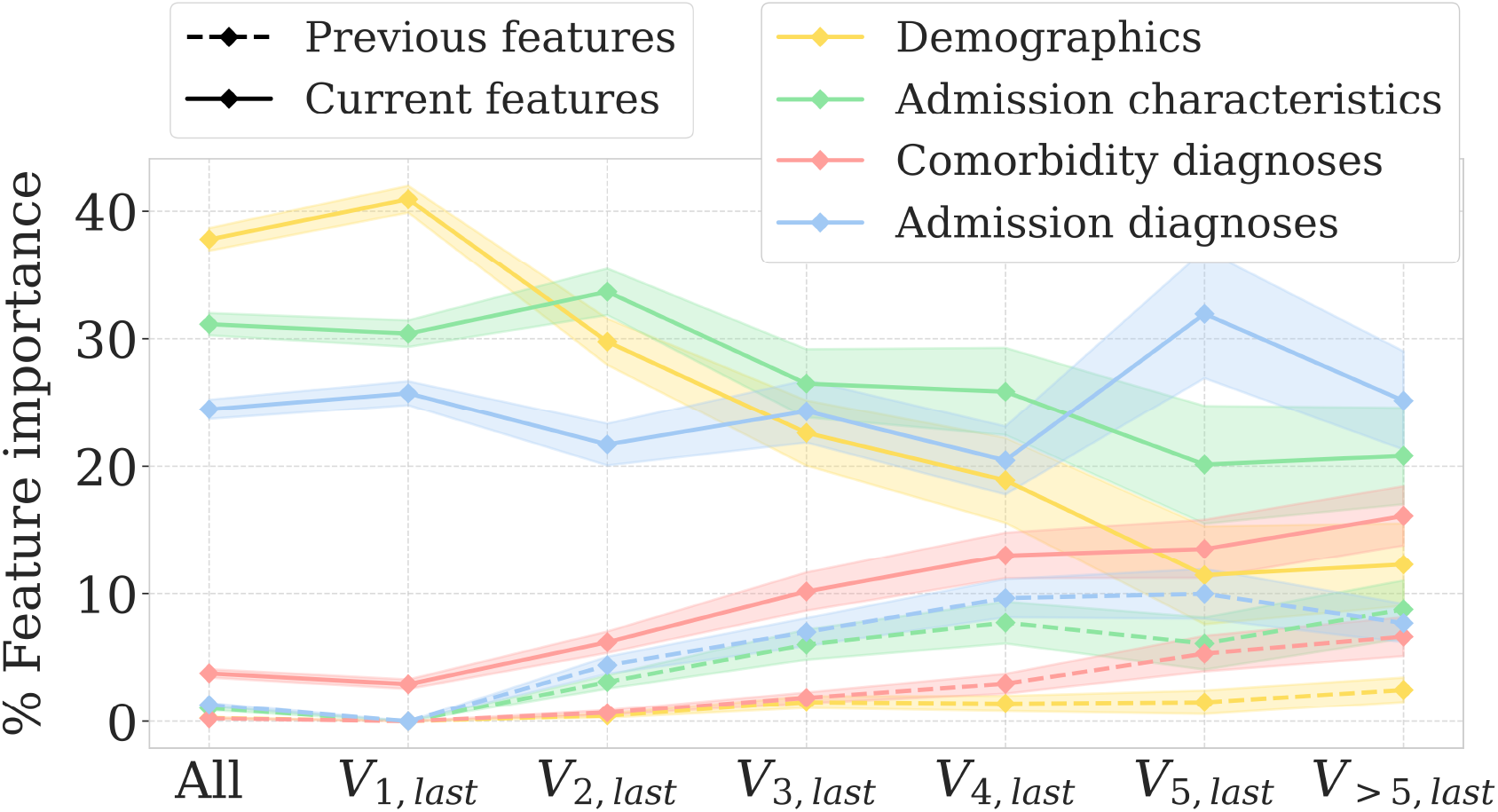
Post-hoc analyses of feature importance of the ELSTM trained with AdmDemoDx predictors when including the time gap between current and previous admissions as a predictor. Importance of each feature is computed using feature permutation [26] over 100 bootstraps. Shaded regions indicate variations within one standard deviation of the mean over 100 bootstraps

We also evaluated the performance of the final ELSTM on various population subgroups within the holdout set. The model performed similarly across patients of different biological sexes (Table 11), but its accuracy varied among age subgroups, with the oldest patients being the least accurately predicted (Table 12).

**Table 11:**
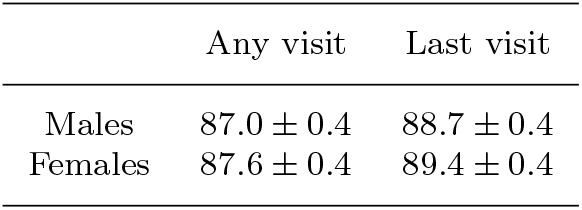
Performance of the ELSTM on subpopulations of males and females. The scores correspond to the *mean ± standard deviation* of the AUROC over 100 bootstraps.

**Table 12:** Performance of the ELSTM on subpopulations of different age groups. The scores correspond to the *mean ± standard deviation* of the AUROC over 100 bootstraps.

|  | Any visit | Last visit |
| --- | --- | --- |
| Age $\leq 50$ | $89.8 \pm 1.0$ | $91.5 \pm 0.9$ |
| $50 < \text{Age} < 65$ | $88.5 \pm 0.7$ | $90.3 \pm 0.6$ |
| Age $\geq 65$ | $82.8 \pm 0.4$ | $84.9 \pm 0.4$ |

## Footnotes

1 https://pypi.org/project/skranger/

2 https://www.octopize.io

3 https://pypi.org/project/skranger/

